# New lesion formation is associated with accelerated brain aging in multiple sclerosis

**DOI:** 10.64898/2026.08.27.26361556

**Authors:** Francesco La Rosa, Jonadab Dos Santos Silva, Emma Dereskewicz, Kamso Onyemeh, Batuhan Ayci, Edward Sizer, Elizabeth Shashkova, Nadia Garcia, Robin Graney, Sarah Levy, Ilana Katz Sand, James Sumowski, Erin S. Beck

## Abstract

**Background:** Brain age is a biomarker of brain tissue integrity associated with disability in multiple sclerosis. While new lesion formation is central to MS diagnosis and treatment monitoring, its direct relationship to brain aging has not been established.

**Methods:** We analyzed 163 people with MS with clinical and MRI assessments at baseline and years 3, 6, and 8. Brain age was estimated using BrainAgeNeXt. Annualized brain age acceleration was modeled as a function of radiological activity using generalized estimating equations, adjusting for age, sex, disease duration, baseline T2 lesion volume, normalized brain volume (NBV), brain age difference (BAD), and disease-modifying therapy. Secondary analyses examined dose-response effects, post-activity recovery, paramagnetic rim lesion (PRL) associations, and disability associations.

**Results:** 105 participants had at least one new T2 lesion over 8 years. Radiologically active intervals (138 of 333) were associated with +0.19 yr/yr greater brain age acceleration than stable intervals (95% CI: 0.03–0.37; p=0.022), scaling with lesion count (β=+0.18; p=0.001) and volume. Older age, greater baseline BAD, and NBV were independently associated with reduced brain age acceleration. Brain age acceleration in individuals with new lesions normalized during subsequent stable intervals (0.41 vs −0.06 yr/yr; p=0.001). Both PRLs and non-PRL lesions were associated with greater brain age acceleration than stable intervals. Baseline BAD, but not annualized acceleration, predicted Expanded Disability Status Scale (EDSS) and Nine-Hole Peg Test (9HPT) worsening.

**Conclusions:** New focal lesion formation is associated with a quantifiable, dose-response acceleration of brain aging in MS that normalizes once lesion activity is suppressed.

## INTRODUCTION

Multiple sclerosis (MS) is an immune-mediated disease characterized by inflammatory demyelination and progressive neurodegeneration, leading to physical and cognitive impairment^1^. White matter lesions, visible on magnetic resonance imaging (MRI), are common in MS and are essential for MS diagnosis and assessment of treatment efficacy^2^. MS, however, is characterized by a dissociation between its focal radiological signature and disability and disability accrual over time^3^. This dissociation, commonly referred to as the clinico-radiological paradox, reflects the limited ability of conventional lesion-based metrics to capture the full extent of ongoing neurodegeneration and emphasizes the importance of assessment of damage in normal-appearing brain tissue.

The impact of a focal white matter lesion is not confined to its radiological borders, as axonal transection triggers Wallerian degeneration that damages distant and structurally connected tissue^4^. Each new lesion, therefore, represents a systemic event that amplifies global neurodegeneration, yet the tools routinely used in clinical practice remain insufficient to quantify this effect. Conventional MRI metrics such as T2 lesion volume (T2LV) and clinical scales such as the Expanded Disability Status Scale (EDSS) lack the sensitivity to detect these diffuse changes, creating a critical gap in our ability to monitor subtle progression in the disease trajectory. Furthermore, while normalized brain volume is a common endpoint in clinical trials to capture overall brain atrophy, it often fails to reflect regional patterns of tissue loss that have been shown to be the most clinically meaningful^5,6^.

Chronological age is the single strongest predictor of clinical course in MS, with older age at onset associated with faster accumulation of irreversible disability^7^. This relationship suggests that aging-related processes, including immunosenescence and reduced CNS repair capacity, contribute to the neurodegenerative burden of the disease independently of focal inflammatory activity^8,9^. Brain age is an emerging marker of brain health and integrity and has emerged as a promising approach to estimate both local and global tissue loss associated with aging^10^. Brain age is most commonly predicted from T1-weighted structural MRI scans using advanced machine learning methods. These models predict an individual’s brain age from high-dimensional features directly extracted from the image^8^. The difference between estimated brain age and chronological age, referred to as Brain Age Difference (BAD), provides a measure of structural decline that is sensitive to neurodegeneration. In MS, brain age is associated with physical disability measures, cognitive performance, and disease progression^9,11^.

In our previous work, we have developed BrainAgeNeXt^9^, a convolutional neural network that predicts brain age from a T1w MRI. BrainAgeNeXt was evaluated in two cohorts of people with MS (pwMS), including the *Reserve against Disability in Early MS* (RADIEMS) study at Mount Sinai^12^. In RADIEMS, pwMS showed elevated brain age, and their BAD was associated with baseline disability measures and structural volumetric markers such as normalized brain volume.

Evaluating lesion-filling on RADIEMS baseline data confirmed that the presence or filling of focal T1 lesions does not confound BrainAgeNeXt’s predictions. Over a 3-year follow-up time, the study revealed that pwMS with clinical disability progression experienced significantly faster annual brain age acceleration than those who remained clinically stable^9^. However, whether focal inflammatory activity is associated with acceleration of brain aging, and the factors that might modulate this relationship remain poorly understood.

In this study, we analyzed 8-year longitudinal data from pwMS in the RADIEMS cohort to investigate the relationship between clinico-radiological factors and brain age acceleration. Using a population-averaged non-linear statistical framework based on generalized estimating equations, we investigated three questions: (i) whether focal inflammatory activity is associated with acceleration of brain aging; (ii) whether this effect follows a dose-response relationship with new lesion number and volume; and (iii) whether baseline normalized brain volume, a measure of structural reserve, is associated with accelerated aging.

## METHODS

### Participants

The RADIEMS cohort is a prospective longitudinal study of 185 people with MS followed at Mount Sinai^12^. The study was approved by the Institutional Review Board (IRB number: 16-00455), and written informed consent was obtained from all participants. 163 subjects with at least one follow-up MRI available were included in this study. Participants were recruited within 5 years of MS diagnosis and underwent a clinical assessment and 3T brain MRI at years 0, 3, 6, and 8 (Y□, Y□, Y□, Y□).

The MRI protocol consisted of 3D Fluid-Attenuated Inversion Recovery (FLAIR) and T1-weighted Magnetization Prepared - RApid Gradient Echo (MPRAGE) at Y0, Y3, Y6, and Y8, 3D T2*-weighted Echo Planar Imaging (EPI) at Y6 and Y8, and T1-weighted Magnetization Prepared - 2 RApid Gradient Echo (MP2RAGE) at Y6 (full acquisition parameters in Supplementary Material, Table 1). Only 60 participants were imaged with MPRAGE at Y6; individuals who instead underwent MP2RAGE at this timepoint were not included in the current analysis. Clinical assessments included Expanded Disability Status Scale (EDSS), Symbol Digit Modalities Test (SDMT), Nine-Hole Peg Test (9HPT), and Timed 25-Foot Walk (T25FW).

**Table 1:**
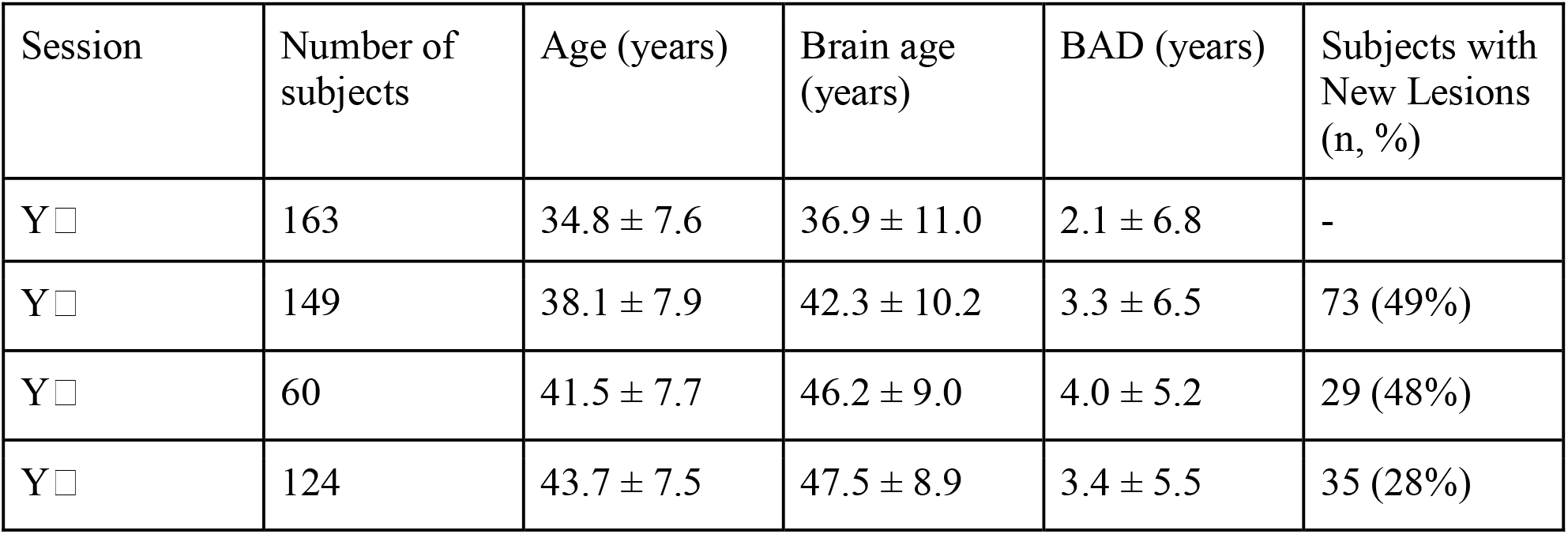
Summary of the cohort characteristics and brain age results. BAD: brain age difference.

| Session | Number of subjects | Age (years) | Brain age (years) | BAD (years) | Subjects with New Lesions (n, %) |
| --- | --- | --- | --- | --- | --- |
| Y<sub>0</sub> | 163 | $34.8 \pm 7.6$ | $36.9 \pm 11.0$ | $2.1 \pm 6.8$ | - |
| Y<sub>1</sub> | 149 | $38.1 \pm 7.9$ | $42.3 \pm 10.2$ | $3.3 \pm 6.5$ | 73 (49%) |
| Y<sub>2</sub> | 60 | $41.5 \pm 7.7$ | $46.2 \pm 9.0$ | $4.0 \pm 5.2$ | 29 (48%) |
| Y<sub>3</sub> | 124 | $43.7 \pm 7.5$ | $47.5 \pm 8.9$ | $3.4 \pm 5.5$ | 35 (28%) |

### Image processing and annotation

MS lesions were segmented on 3D FLAIR using FLAMeS^13^ followed by manual adjustment. New lesions were manually identified by expert raters comparing baseline and follow-up FLAIR and T1w images. On Y_6_ MRIs, PRLs were identified manually by two raters on filtered, unwrapped 3D EPI phase images^14^, followed by consensus review, per published guidelines^15^. Lesions that were new at Y_3_ were assessed for paramagnetic rims on Y_6_ images, and lesions that were new at Y_6_ were assessed for rims on Y_8_ images. Lesion filling was performed with FSL^16^ on T1w images, followed by manual review. Freesurfer 7.4^17^ *recon-all* pipeline was run on the lesion-filled images to compute normalized brain volume (NBV). Percent brain volume change (PBVC) between timepoints was computed using SIENA^16^ on T1w images.

### Brain Age Prediction

Brain age was estimated using BrainAgeNeXt^9^, a convolutional neural network inspired by the MedNeXt framework that predicts brain age directly from minimally preprocessed 3D T1-weighted MRI. All scans underwent skull-stripping with SynthStrip^18^, N4 bias field inhomogeneity correction, and linear registration to the MNI152 1mm isotropic template space using ANTs^19^ prior to inference. Brain age was predicted as the median output across five independently trained ensemble models. The known regression-to-the-mean bias was corrected using the linear correction proposed by Beheshti et al.^20^:

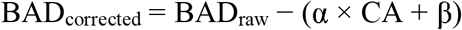

where parameters α = 0.069 and β = −3.544 were estimated from the normative validation set. All subsequent analyses used bias-corrected BAD values.

For each scan-to-scan interval [s1, s2], the annualized brain age acceleration was defined as (BA_s2_ − BA_s1_) / Δyears, where Δyears is the chronological time elapsed between the two scans. A positive value indicates brain aging outpacing chronological aging. Intervals were categorized as radiologically active (≥1 new T2 lesion) or stable (no new lesions).

### Statistical analysis

#### Primary model

Generalized estimating equations (GEE) with an exchangeable correlation structure were used to estimate population-averaged effects of focal inflammatory activity on brain age acceleration. GEE was selected for its robustness to correlation structure misspecification and validity under unbalanced follow-up (1–3 intervals per subject). The model included binary timing group (radiologically active vs stable), chronological age modeled as cubic B-splines (df=3) to capture non-linear aging, baseline BAD at interval start, log-transformed baseline T2 lesion volume, baseline normalized brain volume (NBV), sex, disease duration, and disease-modifying therapies (DMT) efficacy. DMT efficacy was divided into three categories: untreated or low-efficacy DMT, moderate-efficacy DMT, and high-efficacy DMT (see Supplementary Material Table 3 for the full list of DMTs). A similar analysis was repeated with annualized T1w SIENA PBVC in place of brain age acceleration as the outcome.

#### New lesion dose-response model

A secondary GEE model replaced the binary timing indicator with log(1 + new lesion count), a log-transformation accounting for the right-skewed distribution of lesion counts and modeling potential saturation at high lesion burdens. All other covariates were retained.

#### Sensitivity analysis

A Linear Mixed-Effects Model (LMM) with the same fixed-effects specification and per-subject random intercepts was fitted using restricted maximal likelihood to confirm that the effect persisted after accounting for individual heterogeneity in baseline aging and susceptibility to lesion-associated acceleration.

#### Recovery analysis

In subjects with at least one active interval followed by at least one stable interval (n=53), the mean acceleration during active intervals was compared to the mean during subsequent stable intervals using a paired two-sided *t*-test.

#### Paramagnetic rim lesion analysis

We tested whether PRL burden was independently associated with baseline BAD using ordinary least squares regression:

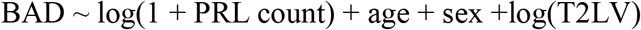

In the longitudinal interval dataset, intervals were classified as radiologically stable, active PRL−, or active PRL+ based on the presence of new T2 lesions with or without a paramagnetic rim. Group differences in annualized brain age acceleration were assessed with a Kruskal-Wallis test, followed by pairwise Mann-Whitney U tests with Benjamini-Hochberg false discovery rate (FDR) correction.

#### Disability associations

Associations between baseline brain age and subsequent disability worsening (for each of the four disability measures: EDSS, SDMT, 9HPT, T25FW) were assessed using linear regression, adjusting for age, sex, and baseline disability score.

Associations between annualized brain age acceleration and disability change over follow-up were assessed using Pearson correlation. To account for multiple comparisons across the four outcomes, p-values were FDR-corrected (p_FDR_), applied separately within each set of four tests.

All analyses were performed in Python 3.10 using statsmodels (v0.14). Statistical significance was set at α=0.05 (two-tailed).

## RESULTS

### Cohort characteristics

Of the original RADIEMS cohort (n=185), 163 people with MS who had at least one follow-up MRI were included (111 females, 68%; 151 relapsing-remitting MS (RRMS), 93%; 12 clinically isolated syndrome (CIS), 7%; mean age 34.8 ± 7.6 years; mean time since MS diagnosis 2.2 ± years, median T2 lesion volume 2171 mm^3^ [IQR 737-5413], median EDSS 1.0 [IQR 0.5-2]). A total of 333 scan-to-scan intervals were analyzed (mean total follow-up time 7.4 ± 1.9 years). Of these, 138 (41%) were classified as radiologically active and 195 (59%) as radiologically stable (Table 1). Mean interval time was 3.2 ± 0.4 years between baseline and Y□, 3.2 ± 0.6 years between YL and Y□, and 1.9 ± 0.5 years between Y□ and Y□. At study entry, mean BAD was +2.1 ± 6.8 years, and was higher at subsequent timepoints vs baseline (+3.3 ± 6.5 years at Y□, +4.0 ± 5.2 years at Y□, +3.4 ± 5.5 years at Y□; all p_FDR_<0.001 vs. Y□; Figure 1), but not between Y□ and the subsequent timepoints. The proportion of active intervals declined from 49% between Y□ and Y□ to 28% between Y□ and Y□, consistent with progressive uptake of high-efficacy DMTs over the study period (Supplementary Material, Table 3).

**Figure 1.**
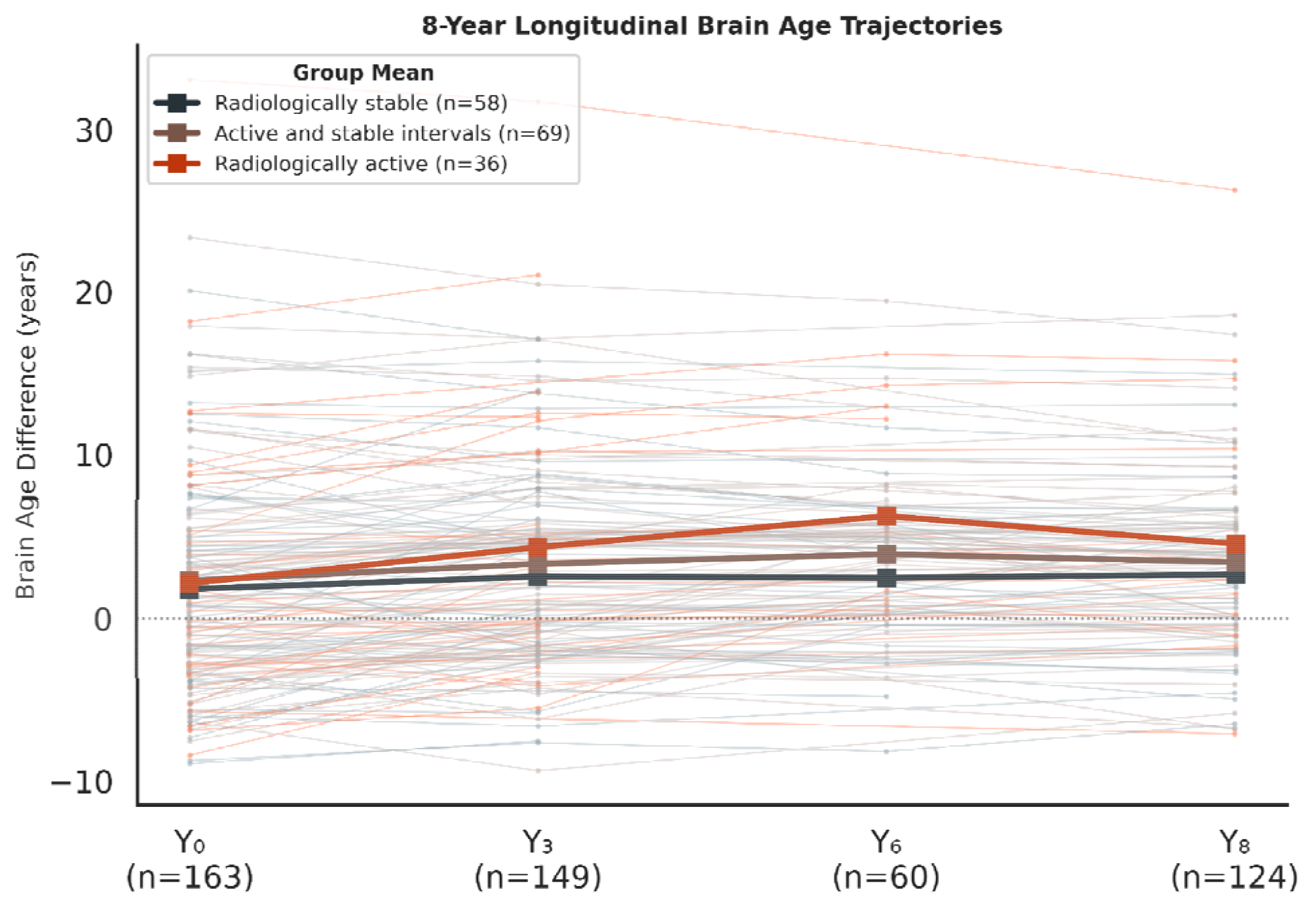
Brain age difference trajectories across timepoints. Individual longitudinal trajectories are shown in light background lines, grouped by lesion-activity pattern across all available follow-up intervals: radiologically active (orange, new lesion(s) at every interval, n=36), active and stable intervals (brown, new lesion(s) at some but not all intervals, n=69), and radiologically stable (gray, no new lesions at any interval, n=58). Bold lines show each group’s mean Brain Age Difference at each timepoint.

### Baseline characteristics and longitudinal brain aging

BAD at the start of each interval was inversely associated with subsequent annualized brain age acceleration (r=−0.293; p<0.001). Younger age was also associated with greater subsequent acceleration in the primary GEE model (age modelled as a cubic B-spline; β=−0.22 per decade, 95% CI −0.39 to −0.05, p=0.013; joint Wald test across spline terms, p<0.001), though this association weakened substantially in the LMM (joint Wald test across spline terms, p=0.048). Baseline NBV was a significant negative predictor of subsequent acceleration in the primary GEE model (β=−0.086 per SD; 95% CI −0.150 to −0.021; p=0.010), and consistent in direction in the LMM (β=−0.096; p=0.057); baseline BAD and NBV were themselves inversely correlated after adjustment for age and sex (partial r=−0.451; p<0.001). In contrast, several baseline factors were not associated with longitudinal brain age acceleration: sex, baseline T2 lesion volume, time since MS diagnosis, and DMT efficacy (all p > 0.05, Figure 2).

**Figure 2.**
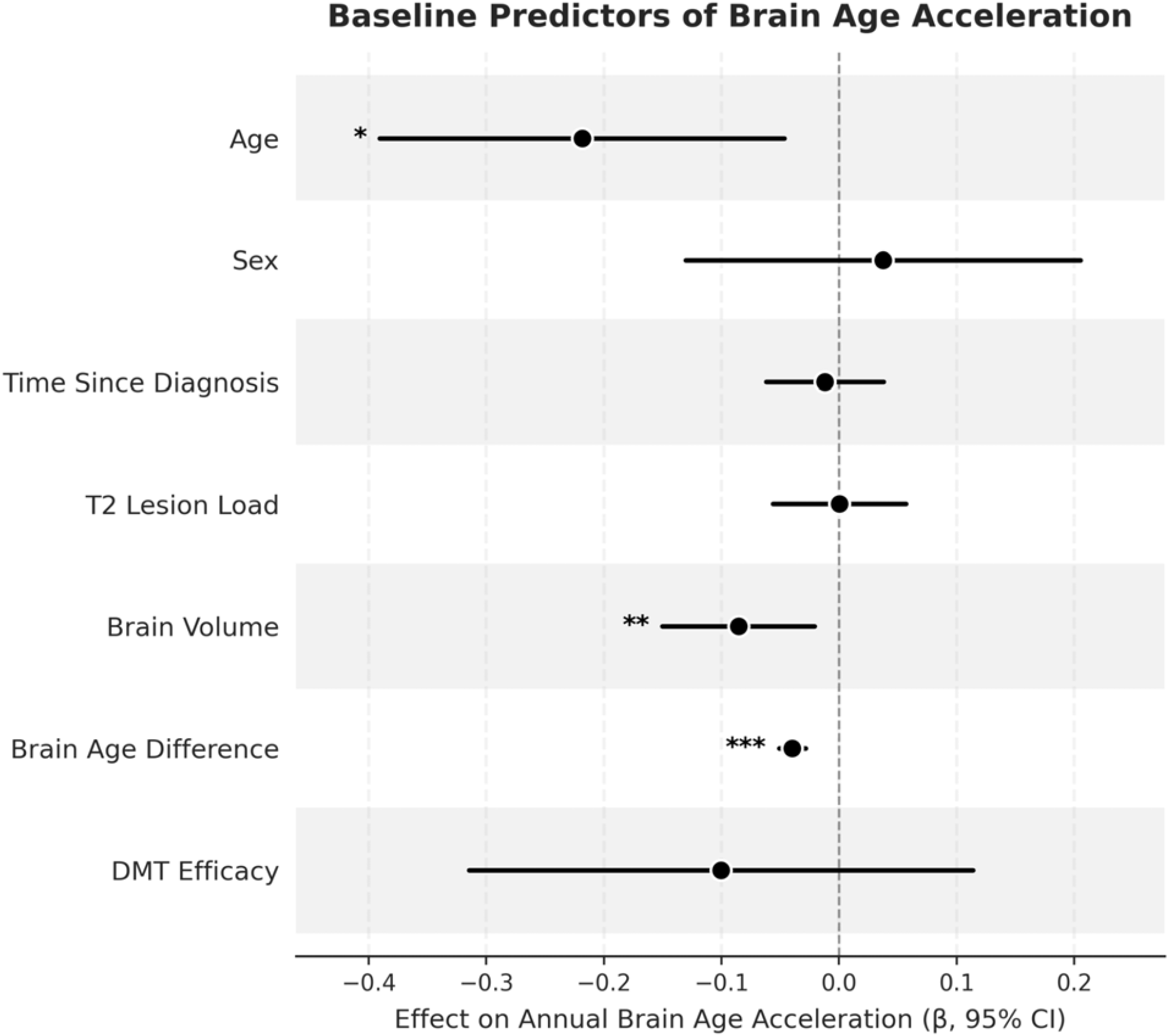
Age, baseline brain age difference, and baseline brain volume are associated with longitudinal brain age acceleration. Forest plot of predictors of annualized brain age acceleration from the generalised estimating equation (GEE) model, adjusted for age (modelled as a cubic B-spline), sex, time since diagnosis, baseline T2 lesion load, baseline brain volume (NBV), and DMT efficacy. Because age was modelled non-linearly, its bar reflects the model-predicted difference in acceleration per decade of age, computed as a proportional rescaling of the contrast between the youngest and oldest participants in the cohort (23–58 years) on the fitted spline. Sex, time since diagnosis, T2 lesion load, and DMT efficacy were not associated with brain age acceleration. New lesion formation was also a significant predictor in this model and is discussed separately below.

### New lesion formation is associated with accelerated brain aging

In the primary GEE model, radiologically active intervals were associated with +0.19 yr/yr greater annualized brain age acceleration than stable intervals (95% CI: 0.03–0.37; p=0.022). This effect was independent of age, sex, time since diagnosis, baseline T2 lesion volume, NBV, and DMT efficacy. Raw distributions of annualized acceleration for stable and active intervals are shown in Figure 3. In the LMM sensitivity analysis, the fixed effect for lesion activity was β=+0.27 yr/yr (95% CI: 0.09–0.44; p=0.003), consistent with the GEE estimate. The same binary comparison for annualized T1w SIENA PBVC was not significant (GEE β=−0.025 %/yr, p=0.44; LMM β=−0.01 %/yr, p=0.74), despite trending in the same direction.

**Figure 3.**
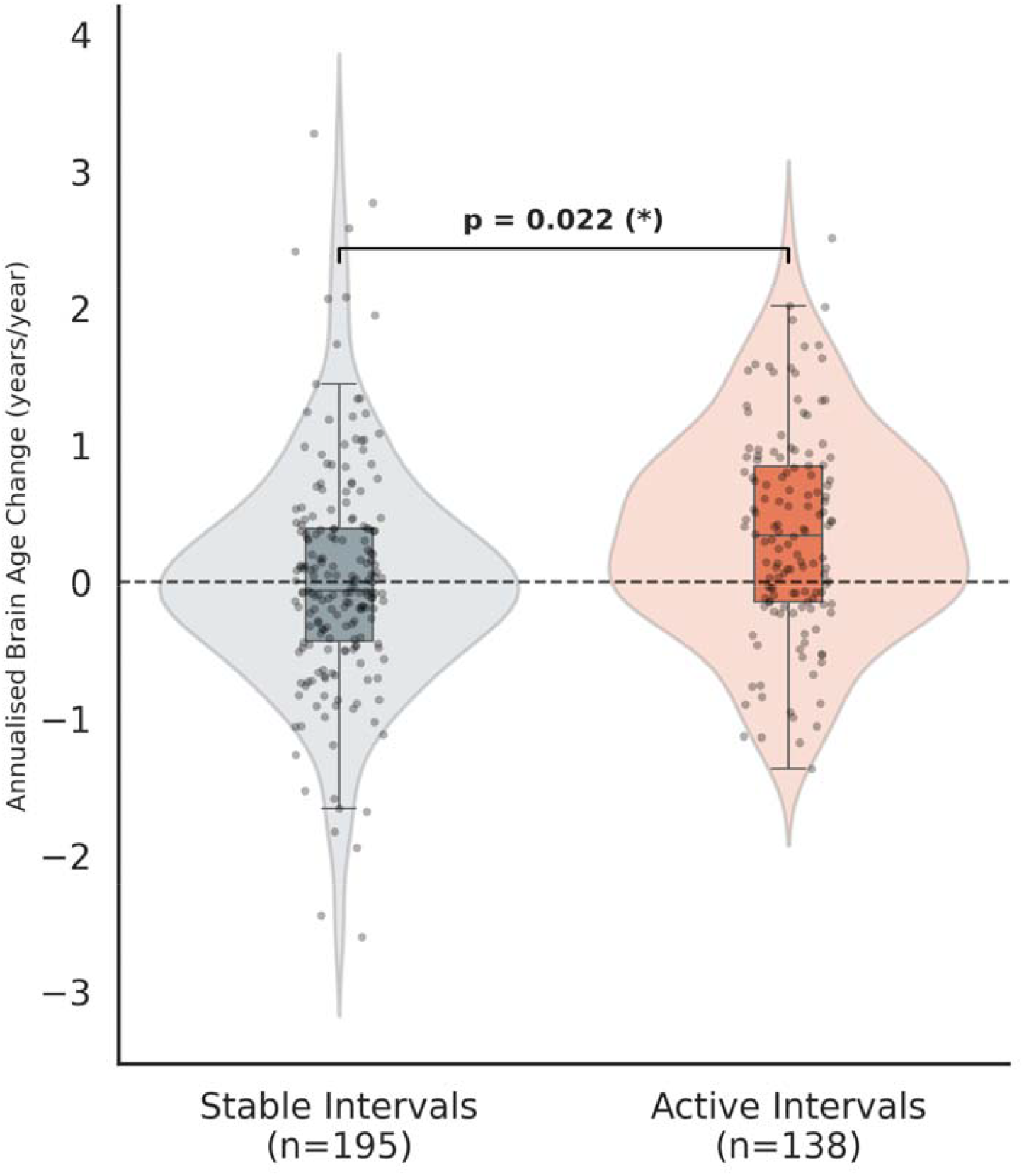
Radiologically active intervals show greater annualized brain age acceleration compared to stable intervals. Violin plot of the annualized brain age acceleration for stable (n=195) and radiologically active (n=138) scan intervals, showing raw, unadjusted data. The p-value shown is from the primary, covariate-adjusted generalised estimating equation (GEE) model (adjusting for age, sex, baseline BAD, T2 lesion volume, NBV, DMT efficacy, and disease duration)

### The number and volume of new lesions are related to the magnitude of brain age acceleration

Replacing the binary new lesion variable with log(1 + new lesion count) confirmed a significant positive dose-response association (GEE β=+0.184; 95% CI: 0.073–0.295; p=0.001; Figure 4). A parallel model using log(1 + new lesion volume) in place of count also showed a consistent significant association (GEE β=+0.048; 95% CI: 0.014–0.083; p=0.006). The same dose-response model for annualized T1w SIENA PBVC also showed a significant association with both new lesion count (GEE β=−0.056 %/yr; 95% CI: −0.093 to −0.019; p=0.003) and volume.

**Figure 4.**
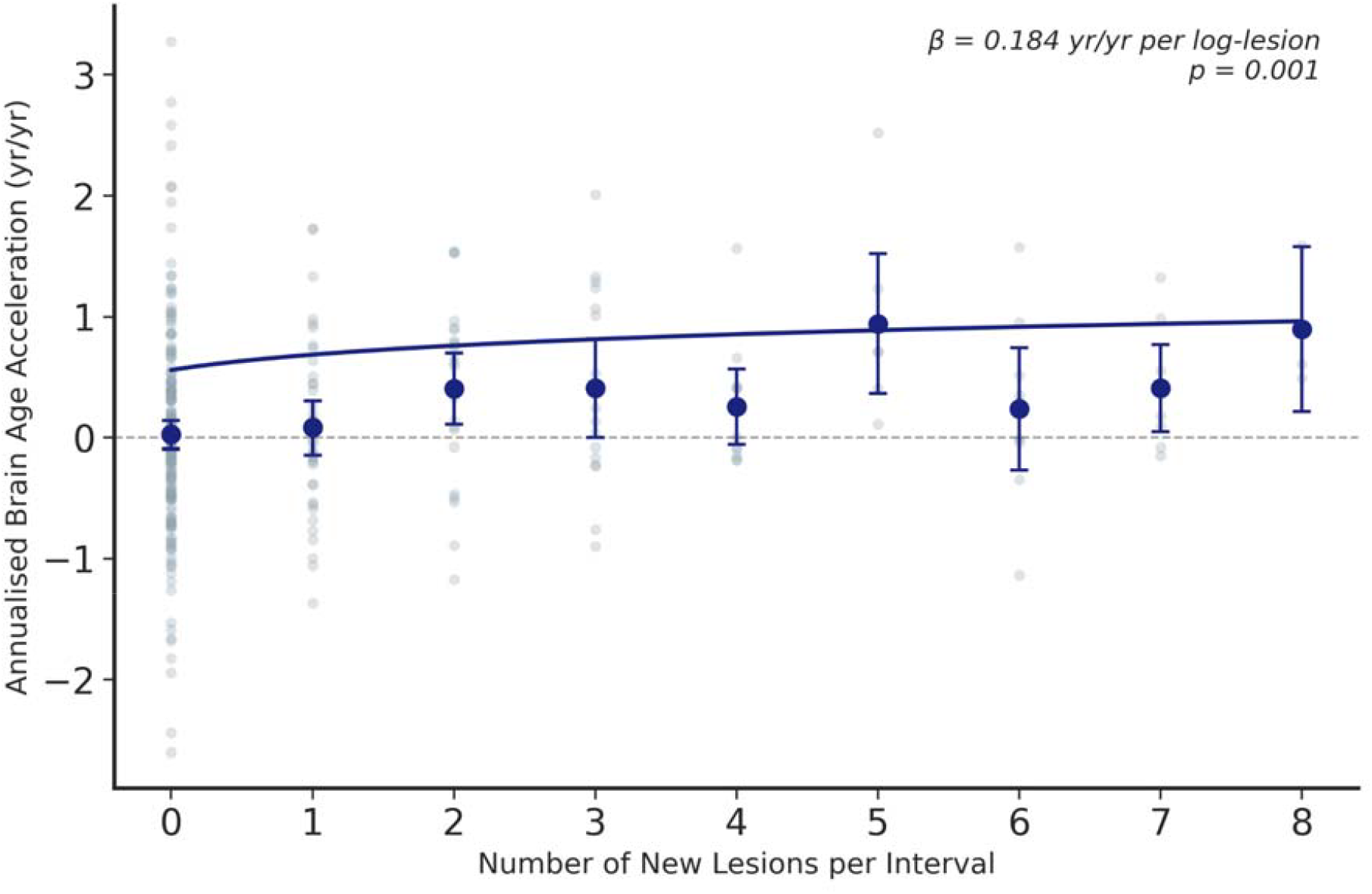
Annualized brain age acceleration increases with the number of new lesions. Dose-response relationship between new lesion number and annualized brain age acceleration. Gray points represent individual scan-to-scan intervals. The fitted curve reflects the generalized estimating equation model, in which new lesion count was log-transformed.

### Brain age acceleration normalizes after lesion control

In individuals with an interval with new lesion formation followed by a stable interval (n=53), mean annualized brain age acceleration during active intervals was +0.41 ± 0.79 yr/yr, compared to −0.06 ± 0.50 yr/yr during subsequent stable intervals (paired *t*-test p=0.001, Cohen’s δ = 0.50; Figure 5). Brain age acceleration during stable intervals was not significantly different from zero (p=0.37). This normalization was not due to treatment escalation: in a subset of 22 subjects who remained on the same DMT efficacy category throughout both periods, the same pattern held (+0.46 ± 0.73 vs. −0.11 ± 0.47 yr/yr; paired t-test p=0.006, Cohen’s δ = 0.65).

**Figure 5.**
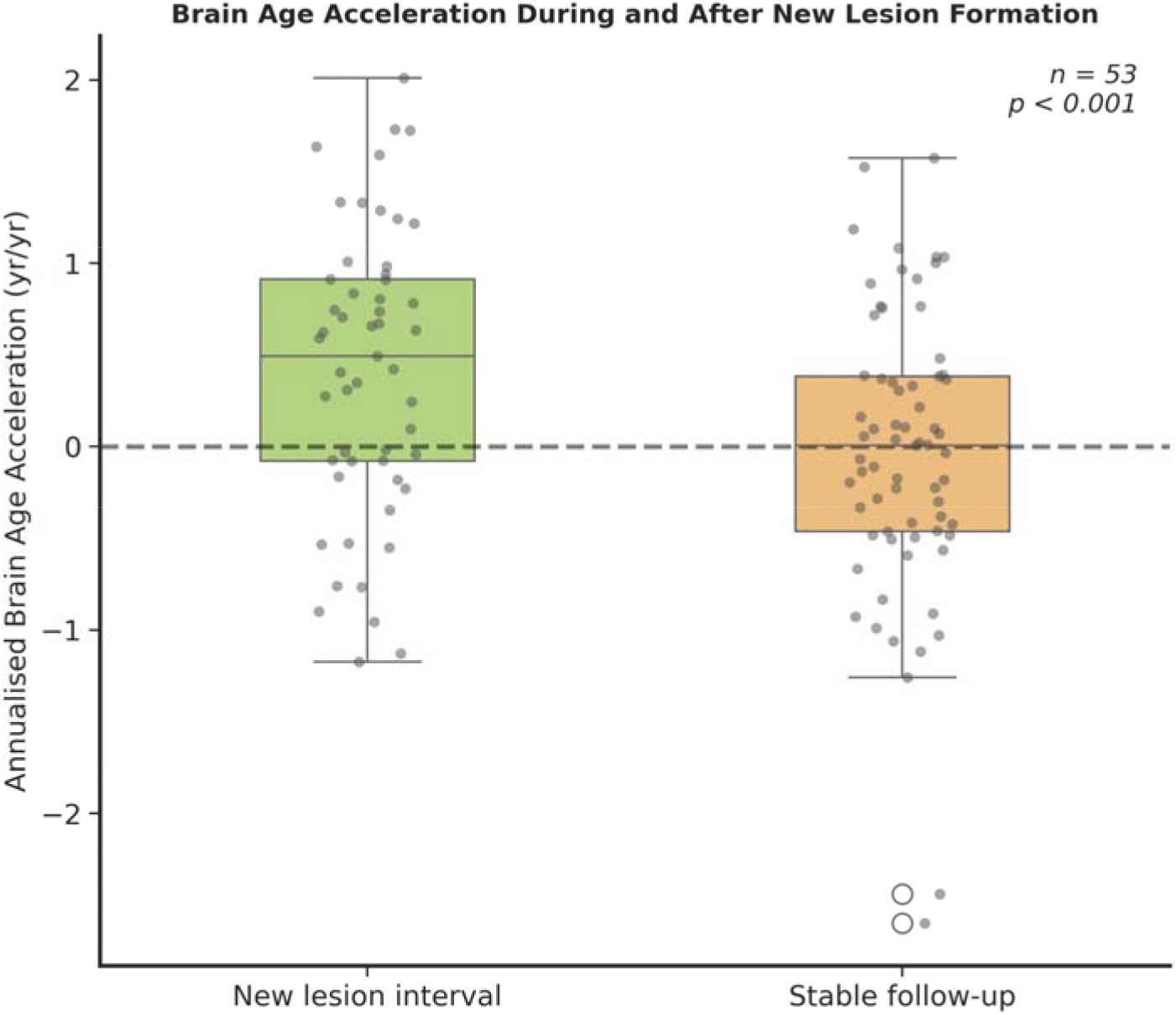
Brain aging rates normalize following control of new lesion formation. Within-subject brain age acceleration during intervals with new lesion formation versus subsequent radiologically stable follow-up intervals. Paired *t*-test compares the active and stable intervals within the same subject (p=0.001).

### Paramagnetic rim lesions and brain aging

Out of 123 participants with a T2*w EPI image available for PRL assessment, 55 (45%) had at least one PRL. People with ≥1 PRL (PRL+) had higher baseline BAD than PRL− participants (4.5 ± 7.9 vs 0.61 ± 4.8 years, p=0.001). After adjusting for age, sex, and T2 lesion volume, PRL count was not independently associated with baseline BAD (p=0.056). Baseline T2 lesion volume was the sole significant predictor in this model (β=+2.04, p<0.001). Further, we define as active PRL+ the intervals in which at least one new PRL formed and active PRL- the ones in which all new lesions were PRL-. Longitudinally, in the subset of intervals available (stable n=165, active PRL− n=90, active PRL+ n=17), both active PRL- and active PRL+ intervals were associated with significantly greater brain age acceleration than stable intervals (PRL−: mean +0.26 yr/yr, p_FDR_=0.003; PRL+: mean +0.54 yr/yr, p_FDR_=0.015; Kruskal-Wallis H=15.2, p<0.001). The difference between PRL− and PRL+ intervals was not significant (p_FDR_=0.14).

### Longitudinal disability associations

Disability remained largely stable at the group level across the 8-year follow-up: EDSS, 9HPT, and T25FW showed no significant group level change, whereas SDMT showed a minimal improvement (Supplementary Material Table 2). Baseline BAD was associated with EDSS (β=+0.048 per year of BAD, p_FDR_=0.001) and 9HPT worsening (β=+0.173 per year of BAD, p_FDR_<0.001) over time, but not with SDMT or T25FW, after adjusting for age, sex, and baseline disability. Annualized brain age acceleration was not significantly associated with change in any disability measure over the 8-year follow-up (all |r|<0.18; all p_FDR_>0.10).

## DISCUSSION

In this eight-year longitudinal study of 163 people with early MS, new focal white matter lesion formation was associated with a period of accelerated brain aging, independently of sex, age, disease duration, DMT efficacy, and baseline lesion burden. The effect followed a dose-response gradient with new lesion count and volume and normalized in subsequent radiologically stable intervals. These findings extend and contextualize our prior BrainAgeNeXt work^9^, which demonstrated that brain age increases in MS over time and that disability worsening is associated with higher brain age accumulation^9^. In this work, we identify a within-subject, interval-level modulator of that excess: the formation of new MRI-visible focal lesions, rather than baseline lesion load or disease duration. These results may be related to the known downstream consequences of each new lesion: axonal transection, Wallerian degeneration cascades, and microstructural damage within normal-appearing tissue.

The magnitude of the brain aging acceleration effect is relevant. A penalty of 0.19 yr/yr during active intervals implies that an individual with five years of active disease accumulates approximately one additional biological year of brain aging beyond chronological time. Given that BAD is already elevated by over two years in our early MS cohort at baseline, consistent with prior evidence of accelerated aging from the earliest stages of MS^9,21,22^, this compounding effect suggests that lesion activity could substantially advance the neurodegenerative trajectory even in young, mildly disabled individuals. The association between new lesions and accelerated brain aging scales proportionally with both the number and volume of new lesions, and the log relationship is consistent with sub-linear saturation at high lesion counts. These findings support the hypothesis that each lesion may contribute independently to more widespread structural brain changes. This dose-response pattern was corroborated using PBVC measured with SIENA. While a single new lesion was not associated with accelerated PBVC compared to radiologically stable intervals, PBVC showed the same significant dose-response relationship with new lesion count as brain age did, in accordance with previous studies linking new lesion formation to greater subsequent brain atrophy^23,24^. This suggests that brain age may be more sensitive to the effect of isolated lesions, while both measures converge once lesion burden accumulates. The sub-linear relationship may reflect saturation of vulnerable axons within connected tracts, or compensatory neuroplasticity at high lesion burden.

Individuals with greater brain volume showed a lower brain age acceleration, independent of lesion activity. The negative correlation between baseline BAD and NBV is expected and has been shown previously^10^, as both represent a measure of brain health and capture brain atrophy. The association between NBV and lower brain age acceleration aligns with the concept of brain structural reserve as a buffer against disease-related neurodegeneration^25^, but could also indicate that prior brain age acceleration/brain atrophy is associated with continued brain age acceleration over time.

The recovery analysis provides the most actionable finding: brain age acceleration normalized once lesion activity was suppressed, even in subjects who stayed in the same DMT category. This suggests that a component of the effect reflects ongoing, dynamically suppressible injury rather than purely irreversible neurodegeneration^26^. Younger age was independently associated with greater brain age acceleration (Figure 2). This may partly reflect a ceiling effect: participants who are older have less remaining capacity for further deviation of brain tissue integrity from what would be expected at their chronological age, consistent with reports that older individuals with RRMS, while showing lower baseline brain volumes than younger participants, exhibit slower subsequent atrophy rates independent of disease duration^27^. We hypothesize that a similar ceiling dynamic explains why participants with a higher baseline BAD showed less subsequent acceleration, consistent with earlier saturation of the most vulnerable neural tissue in individuals with an already elevated brain age.

Cross-sectionally, participants with PRLs had higher BAD, but this was fully explained by their greater T2 lesion volume. Longitudinally, both new PRL- and PRL+ were associated with accelerated brain aging compared to stable intervals (+0.26 and +0.54 yr/yr respectively). Larger studies will be needed to determine whether chronic active inflammation confers an independent effect above acute lesion formation on brain aging.

When examining the relationships between BAD and disability change over time, baseline BAD predicted subsequent EDSS and 9HPT worsening over follow-up, whereas brain age acceleration itself was not associated with disability change over time. This last pattern may be due to the delay between MRI-detectable neurodegeneration and clinical disability accumulation in MS as well as the very limited longitudinal changes in disability in the RADIEMS cohort (Supplementary Material, Table 2). Longer follow-up will be required to determine whether brain age acceleration predicts future disability worsening.

Several limitations should be acknowledged. Due in part to changes in imaging protocols, brain age was only assessed in only 41 of 163 participants (25%) at all four timepoint visits and only 60/163 Y_6_ scans included MPRAGE, in some cases limiting our ability to assess brain age change temporally associated with new lesion formation with high temporal resolution. In addition, PRLs were only assessed at Y_6_ and only in 123 participants, which may have limited our power to detect associations between brain age acceleration and chronic active inflammation within new or chronic lesions. The cohort is young and mildly disabled, limiting generalizability to progressive MS and older populations. BrainAgeNeXt^9^ was developed to predict brain age from a single T1w MRI and may introduce some measurement noise when applied on longitudinal data, as it was not explicitly trained or validated for scan-rescan consistency across repeated timepoints in the same individual. This could add variability to interval-level acceleration estimates and may have attenuated the true magnitude of the lesion-associated effect. Finally, while temporal ordering is consistent with causality, additional confounders in this observational cohort may be present and a causal relationship between new lesion burden and brain age acceleration cannot be established.

In conclusion, new focal lesion formation is associated with quantifiable acceleration of brain aging in MS that scales with lesion count and volume and largely normalizes once lesion activity is suppressed. This reversibility is consistent with new lesion formation contributing to diffuse structural brain changes, reinforcing the case for early, high-efficacy DMT to prevent any new lesion. Future studies in larger cohorts should investigate other factors that may modulate brain age acceleration, including the effect of high-efficacy DMTs beyond suppressing new lesion formation, and whether brain age trajectory constitutes a valid surrogate endpoint for neuroprotection in clinical trials.

## Data Availability

All data produced in the present study are available upon reasonable request to the authors

## FUNDINGS AND ACKNOWLEGMENTS

This work was also supported by Schmidt Sciences and the Office of the Assistant Secretary of Defense for Health Affairs through the Multiple Sclerosis Research Program under Award No. (HT9425-24-1-0857). Opinions, interpretations, conclusions, and recommendations are those of the author and are not necessarily endorsed by the Department of Defense. Funding was provided by NIH R01HD082176 and R01NS135228. This work was also supported by the computational and data resources and staff expertise provided by Scientific Computing and Data at the Icahn School of Medicine at Mount Sinai and supported by the Clinical and Translational Science Awards (CTSA) grant UL1TR004419 from the National Center for Advancing Translational Sciences. Additional support was provided by the Icahn School of Medicine Capital Campaign, BioMedical Engineering and Imaging Institute, and Department of Radiology. We thank the faculty and staff at the Mount Sinai CGD MS Center for their assistance with participant recruitment and study visits.

