## Supplementary Material for "New lesion formation is associated with accelerated brain aging in multiple sclerosis"

| Sequence | TR (ms) | TE (ms) | TI (ms) | Flip Angle | FOV (mm) | Slices | Voxel size ($mm^3$) | Timepoints |
|---|---|---|---|---|---|---|---|---|
| 3D FLAIR | 5000 | 387 | 1800 | - | 230 | 192 | 0.9x0.45x0.45 | Y0, Y3, Y6, Y8 |
| T1w MPRAGE | 2400 | 2 | 900 | 8° | 256 | 176 | 1.0 (isotropic) | Y0, Y3, Y6, Y8 |
| 3D T2*w EPI | 64 | 35.0 | - | 10° | 256 | 384 | 0.65 (isotropic) | Y6, Y8 |
| T1w MP2RAGE | 5000 | 2.98 | 700 / 2500 (TI1/TI2) | 4° / 5° (α1/α2) | 256 | 192 | 1.0 (isotropic) | Y6 |

**Supplementary Table 1: MRI acquisition parameters.** FLAIR: Fluid-Attenuated Inversion Recovery; MPRAGE: Magnetization Prepared - RApid Gradient Echo; EPI: Echo Planar Imaging; MP2RAGE: Magnetization Prepared - 2 RApid Gradient Echo; TR: repetition time; TE: echo time; TI: inversion time; FOV: field of view. For MP2RAGE, TI and flip angle columns show the two values used for the first and second inversion images (TI1/TI2 and α1/α2, respectively).

| Session | EDSS Median [IQR] | SDMT Mean ± SD | 9HPT Mean ± SD | T25FW Median [IQR] (s) |
|---|---|---|---|---|
| $Y_0$ (n=163) | 1.0 [0.5-2.0] | 57.5 ± 11.1 | 19.4 ± 3.2 | 4.05 [3.76-4.44] |
| $Y_3$ (n=149) | 1.0 [0.0-2.0] | 59.5 ± 12.2 | 19.6 ± 3.8 | 3.96 [3.63-4.38] |
| $Y_6$ (n=60) | 1.0 [0.0-1.5] | 58.5 ± 13.5 | 19.5 ± 3.2 | 3.90 [3.70-4.28] |
| $Y_8$ (n=124) | 1.0 [0.0-2.0] | 60.8 ± 13.4 | 19.7 ± 4.9 | 3.95 [3.72-4.46] |

**Supplementary Table 2: Summary of clinical measures at each timepoint.** EDSS, 9HPT, and T25FW showed no significant change from baseline to last available assessment (all $p>0.10$); SDMT improved significantly ($p<0.001$), likely reflecting a practice effect.

| | None or Low Efficacy DMTs | Moderate Efficacy DMTs | High Efficacy DMTs |
|---|---|---|---|
| List of treatments | No treatment, Interferon Beta 1a, Interferon Beta 1b, Glatiramer acetate, Peginterferon beta-1a | Fingolimod, Ozanimod, Dimethyl fumarate, Diroximel fumarate, Teriflunomide, | Natalizumab, Ocrelizumab, Ofatumumab, Ublituximab, Rituximab, Cladribine |
| $Y_0$ (n=163) | 33 (20%) | 98 (60%) | 32 (20%) |
| $Y_3$ (n=149) | 22 (15%) | 61 (42%) | 63 (43%) |
| $Y_6$ (n=60) | 9 (15%) | 22 (37%) | 28 (47%) |
| $Y_8$ (n=124) | 14 (11%) | 35 (28%) | 75 (61%) |

**Supplementary Table 3: Summary of disease modifying therapies (DMTs).**
